# Estimating the asymptomatic proportion of SARS-CoV-2 infection in the general population: Analysis of a nationwide serosurvey in the Netherlands

**DOI:** 10.1101/2021.03.29.21254334

**Authors:** SA McDonald, F Miura, ERA Vos, M van Boven, H de Melker, F van der Klis, R van Binnendijk, G den Hartog, J Wallinga

**Affiliations:** Centre for Infectious Disease Control, Netherlands National Institute for Public Health and the Environment, Bilthoven, the Netherlands; Center for Marine Environmental Studies (CMES), Ehime University, Matsuyama, Ehime, 790-8577, Japan

**Keywords:** SARS-CoV-2, asymptomatic disease, serosurvey, the Netherlands

## Abstract

**Background:** The proportion of SARS-CoV-2 positive persons who are asymptomatic – and whether this proportion is age-dependent – are still open research questions. Because an unknown proportion of reported symptoms among SARS-CoV-2 positives will be attributable to another infection or affliction, the *observed*, or ‘crude’ proportion without symptoms may underestimate the proportion of persons without symptoms that are *caused* by SARS-CoV-2 infection.

**Methods:** Based on a large population-based serological study comprising test results on seropositivity and self-reported symptom history conducted in April/May 2020 in the Netherlands (*n*=3147), we estimated the proportion of reported symptoms among those persons infected with SARS-CoV-2 that is attributable to this infection, where the set of relevant symptoms fulfills the ECDC case definition of COVID-19, using inferential methods for the attributable risk (AR). Generalised additive regression modelling was used to estimate the age-dependent relative risk (RR) of reported symptoms, and the AR and asymptomatic proportion (AP) were calculated from the fitted RR.

**Results:** Using age-aggregated data, the estimated AP was 70% (95% CI: 65-77%). The estimated AP decreased with age, from 80% (95% CI: 67-100%) for the <20 years age-group, to 55% (95% CI: 48-68%) for the 70+ years age-group.

**Conclusion:** Whereas the ‘crude’ AP represents a lower bound for the proportion of persons infected with SARS-CoV-2 without COVID-19 symptoms, the AP as estimated via an attributable risk approach represents an upper bound. Age-specific AP estimates can inform the implementation of public health actions such as targetted virological testing and therefore enhance containment strategies.

---

The proportion of SARS-CoV-2 positive persons who are asymptomatic is still an open research question [1-4]. Given that age-dependence for symptom occurrence and severity appears plausible [5,6], it will be of considerable public health value to estimate this proportion stratified by age-group; for instance, the efficiency of screening strategies to find asymptomatically infected cases might be improved if adapted according to age.

In this study we estimate the asymptomatic proportion (AP) from a large population-based serosurvey dataset containing IgG antibody (against the spike S1-antigen) test result and history of symptom occurrence as elicited by questionnaire [7]. This dataset presents a unique challenge for estimation and interpretation of the proportion of symptomatic infections due to SARS-CoV-2; participants were unaware of their serostatus and were asked to report any occurrence of symptom(s) from an indicative set of respiratory and related symptoms that they had experienced since the recorded beginning of the outbreak in the Netherlands. An unknown proportion of symptom reports among SARS-CoV-2 seropositive participants will be attributable to another circulating infection, such as the common cold, or seasonal affliction (e.g., hay fever). To use datasets in which reported symptoms could have occurred over a relatively long time-frame, methodology is needed to adjust for symptoms that are *not* associated with SARS-CoV-2 infection.

In the population-based serosurvey data we analyse here, 59% of seropositive respondents reported a symptom history fulfilling the ECDC case definition for COVID-19, but 30% of seronegative respondents also reported eligible symptom(s). Logically, some proportion of the seropositives’ symptoms could be due to another cause, and thus the SARS-CoV-2 attributable percentage would be lower than 59%. Here, we estimate the symptomatic proportion (and its complement, the asymptomatic proportion) through standard methods for estimating the attributable risk (AR).

## METHODS

AR, also termed the *attributable proportion* and the *attributable fraction among the exposed*, estimates the excess risk of disease among exposed persons that can be attributed to exposure. We apply AR methods to estimate the proportion of symptom occurrence among those persons infected with SARS-CoV-2 that is attributable to their infection, where the set of relevant symptoms matches a standard case definition. With the AR approach, one accounts for spurious symptom occurrence in the reporting time-frame, by bringing the risk of symptoms in seronegative persons into the analysis. AR provides an analogue, for observational data, of the difference in risk of symptom occurrence between the exposed (i.e., SARS-CoV-2 seropositive) and unexposed arms of a hypothetical randomised trial.

### Study population

Early during the first wave of COVID-19 in the Netherlands, participants from a large-scale, nationally representative serosurvey (PIENTER-3, conducted in 2016/17 [8]) were invited to participate in the PIENTER-Corona (PICO round 1) follow-up study. The primary aim was to determine population-level SARS-CoV-2 seroprevalence [7]. PICO-1 participants (age range 2-90 years) filled in an online survey form and took a fingerprick blood sample within the period 31 March 2020 through 11 May 2020. The majority (80%) of samples were taken in the first week of April 2020. Further details are provided in Vos et al. [7].

### Case definition

#### Symptomatic cases

The PICO-1 survey queried symptom history since the start of the coronavirus outbreak in the Netherlands (27 February 2020), namely the occurrence of: *fever/chills, general fatigue, cough, sore throat, runny nose, shortness of breath, diarrhea, nausea/vomiting, headache, irritability/confusion, muscle pain, pain while breathing, stomachache, joint pain, loss of smell and taste*, or *other*. Participants were also asked to report the symptom onset and offset dates, estimating these dates if necessary.

A participant was defined as ‘symptomatic’ if they reported the occurrence of symptom(s) matching the ECDC case definition for COVID-19 (fever and/or cough and/or shortness of breath and/or loss of smell/taste) [9]. Because an IgG response takes time to build up and thus be detectable, we *a priori* defined a period of 14 days in which symptom occurrence was discounted [10]. That is, all participants who reported an onset date for symptom(s) that was 0–14 days prior to the survey date were considered ‘not symptomatic’. Among all participants who had reported symptom(s) matching the ECDC case definition (*n*=1172), 324 (28%) did not supply a symptom onset date; these participants were all defined as symptomatic. A small number of participants with reported symptom onset and offset before 27 February 2020 (*n*=11) were classified as not symptomatic. The dataset (*n*=3147) consists of 956 participants who were defined as symptomatic and 2191 defined as not symptomatic (Table S1).

#### Seropositives

Serostatus was determined using a validated immunoassay and was based on a cut-off for antibody concentration while controlling for false positive test results, resulting in high expected specificity of the test [7,11].

### Attributable risk and asymptomatic proportion

AR is defined in terms of the risk in the exposed (R_e_) relative to the risk in unexposed (R_u_): AR = (RR – 1)/ RR. This relative risk (RR) can be estimated using regression analysis methods. We estimate the symptomatic proportion (SP) attributable to SARS-CoV-2 as the proportion of participants in a given age-group who were symptomatic, multiplied by the attributable risk: (R_e_ × AR). The asymptomatic proportion (AP) is then 1 – SP.

### Poisson regression using generalised additive model (GAM) approach

Following Zou [12], a Poisson regression model with robust standard errors was fitted to the outcome variable to estimate the relative risk of symptom(s) associated with SARS-CoV-2 serostatus, while adjusting for the following *a priori* selected covariates: sex, education level [of the mother, if respondent was under 15 years of age; high vs. low/middle], an indicator variable if the respondent [or the parent(s) of respondents <15 years old] works in the healthcare sector, and a categorical variable for the number of people in the household [1, 2, 3 or more]).

As a first step we omitted covariates and fitted penalised splines (via the gam() and s() functions in R package mgcv) to age, separately for seropositive and seronegative persons, as *a priori* we suspected that age would act as a (non-linear) effect modifier of the association between symptom occurrence and serostatus. In subsequent model fits we tested possible covariates, and we selected models with covariates based on the Akaike Information Criterion (AIC).

The RR was estimated, per year of age, by subtracting the linear predictor (on the log scale) for negative serostatus from the linear predictor for positive serostatus. 95% confidence intervals (CIs) were calculated via bootstrapping methods (500 draws with replacement).

Next, AR was calculated using the derived RR. We define the risk of symptoms in the exposed using the predicted value 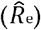 rather than the (sparse) observed value (R_e_). Thus, SP is defined as 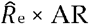, and AP as its complement, 1 – SP. To aid interpretation, we estimated AP for discrete age-groups using the estimated RR for the midpoint of each age-group based on the observed range: 11 for 2-19 years, 25 for 20-29, and similarly for 30-39, 40-49, 50-59, 60-69, and 80 for the oldest (70-90 years) age-group.

### Sensitivity analyses

In sensitivity analysis 1, two alternative durations (10 days and 18 days) of the antibody response time window are investigated, given imprecision in this parameter. In sensitivity analysis 2, ‘symptomatic’ is defined broadly, as the occurrence of *any* symptom from the list in the survey (including ‘other’).

## RESULTS

Fifty-nine percent (44/74) of SARS-CoV-2 seropositives reported symptom(s) matching the ECDC case definition, leading to a crude AP of 41%. Thirty percent (912/3073) of seronegative participants reported symptoms (Table S1). Application of the AR method to age-aggregated data resulted in an estimated AP of 70% (95% CI: 65-77%).

The Poisson GAM regression model yielded a significant effect of positive serostatus (RR of 2.04, 95% CI: 1.68-2.49). Including covariates did not improve model fit (Table S2). Fig. 1 shows the penalized-spline fits to age, plotted separately for seropositive and seronegative participants. For seronegatives, there was a decreasing non-linear risk of symptom(s) with advancing age, but the smooth term for seropositives was not statistically significant.

**Fig. 1.**
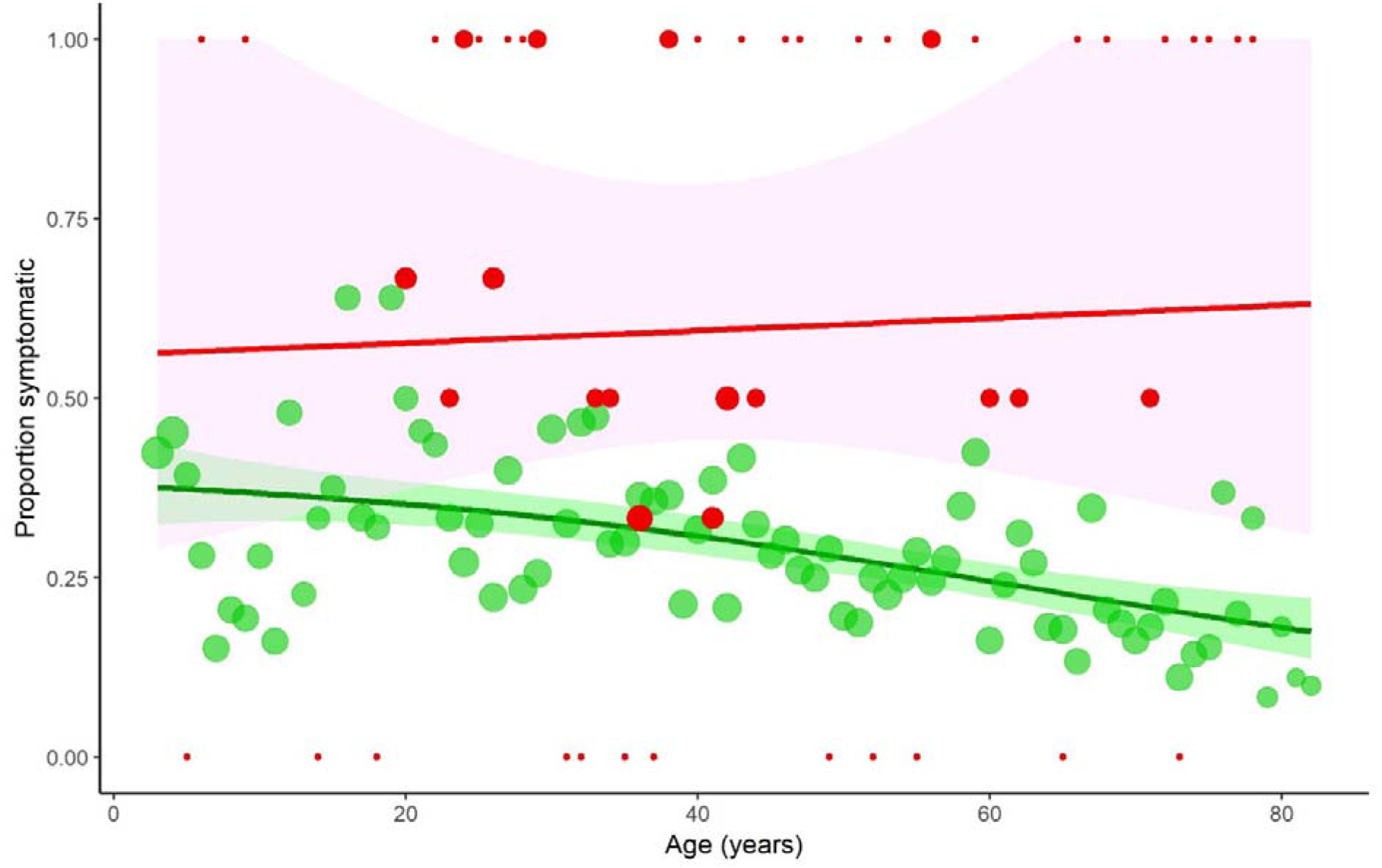
Observed data (as mean proportion symptomatic participants per year of age) and Poisson regression model fit, separately showing the penalised spline fits to seropositive and seronegatives (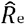and 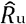, as red and green series, respectively), with 95% confidence bands. Point size indicates the number of data points

The highest AP (approximately 80%) was estimated for the youngest (<20 years) age-group, which declined to about 55% for the oldest (70+ years) age-group (Table 1). The age-dependency in AP is driven by the decreasing frequency of reported symptoms among the seronegatives with age.

**Table 1.** Observed proportion with reported symptoms matching the ECDC case definition, and estimated attributable risk (AR) and asymptomatic proportion (AP), per age-group.

| Age-group | Observed proportion<br>symptomatic (%) [n/N] | Estimated<br>AR (%) (95% CI) | Estimated<br>AP (%) (95% CI) |
| --- | --- | --- | --- |
| <20 years | 50 [4/8] | 35.7 (0.7–58.3) | 79.6 (66.8–99.6) |
| 20-29 | 79 [11/14] | 41.0 (20.4–53.9) | 76.2 (68.7–88.1) |
| 30-39 | 42 [8/19] | 45.6 (27.5–55.8) | 73.1 (67.1–83.8) |
| 40-49 | 64 [7/11] | 50.9 (29.3–60.3) | 69.5 (63.9–82.4) |
| 50-59 | 63 [5/8] | 56.9 (40.3–65.9) | 65.5 (60.0–75.5) |
| 60-69 | 57 [4/7] | 62.9 (45.5–73.9) | 61.2 (54.5–72.0) |
| 70+ years | 71 [5/7] | 71.3 (50.8–83.4) | 55.1 (47.5–68.0) |

The estimated AP was sensitive to the size of the assumed antibody response window; discounting all symptoms reported within a longer, 18-day window prior to data collection resulted in lower estimated ARs and consequent higher APs (Fig. S1). The converse was true when assuming a shorter, 10-day antibody response window.

The results of sensitivity analysis 2 are presented in Table S3 and Fig. S2. Classifying participants as symptomatic if they reported any symptom resulted in higher estimates of the AP compared with using the ECDC case definition. In contrast to the main analysis, in the GAM variable selection two covariates were retained; male sex and low/middle education were associated with a lower reported symptom risk (RRs of 0.90 (95% CI: 0.94-0.97) and 0.91 (95% CI: 0.85-0.98), respectively).

## DISCUSSION

In this study we applied classical AR methods to the problem of estimating the proportion of SARS-CoV-2 seropositive persons whose reported symptom(s) could be attributed to their infection. Using this approach we could estimate both the age-aggregated and age-specific AP, while explicitly accounting for other causes of one or more of the symptoms as reported by seronegatives. The ‘crude’ AP that forms the basis for meta-analytic methods (from which central estimates range from 16 to 23% [2-4]) could underestimate the proportion of persons without symptoms that are caused by SARS-CoV-2 infection, as spurious symptom occurrence – which can be estimated from data on persons testing SARS-CoV-2 negative – is not taken into account.

The difference between ECDC and ‘any symptom’ (sensitivity analysis 2) case definitions in terms of covariate associations suggests that the covariates sex and education level are more related to non-specific symptoms than to symptoms indicative of COVID-19; such symptoms may have been caused by other respiratory viruses, such as influenza and rhinovirus, known to have been in circulation during the symptom reporting time-frame [13].

Estimates were sensitive to the degree of discounting of symptom occurrence close to the sample date. The baseline value of 14 days was a median estimate [10]; we could not account for individual variation in the time needed to build up of an IgG response. Related to this issue, a subset of infected persons (mild/asymptomatic presentations) do not seem to seroconvert at all [14]. This unknown, but likely small degree of misclassification would not have a large impact on our estimate of the AP.

AR quantifies symptom occurrence that is uniquely attributable to SARS-CoV-2 seropositivity. This means that the risk of symptom(s) that co-occur equally frequently with SARS-CoV-2 infection and with another cause (common cold, for example), is ‘removed’ from the AR estimate. AP should be interpreted as the proportion of SARS-CoV-2 infected persons whose symptoms are not attributable to this virus (they may still experience symptoms). The AP is affected by (a) the specificity of the case definition and (b) the time-frame for symptom reporting. The less specific the case definition, the lower the AR and higher the AP (as illustrated in sensitivity analysis 2). Similarly, estimating the AP from survey data relying on symptom recall over a relatively long time-frame will tend to produce lower AR values (and consequently higher APs) compared with a data-elicitation approach that is able to instead measure the occurrence of symptoms that are more directly associated with infection (such as in the window of time several days prior to several weeks following a positive PCR test). For this reason, the AP derived using the AR approach represents an upper bound for the desired parameter: the proportion of persons without symptoms caused by SARS-CoV-2 infection.

In conclusion, classical epidemiological methods for attributing risk of an outcome to exposure may offer a solution for estimating the proportion of SARS-CoV-2 infected persons who are asymptomatic from large-scale surveys measuring serostatus and symptom occurrence. Besides its value for the research and clinical communities [1], age-specific AP can inform the implementation of public health actions, for instance by adapting virological testing strategies according to age to reduce the chance of missing asymptomatic infectives, and therefore enhancing containment strategies.

## Data Availability

Our data are accessible to researchers after publication and upon reasonable request for data sharing to the corresponding author.

