## Supplemental materials for "Estimating the asymptomatic proportion of SARS-CoV-2 infection in the general population: Analysis of a nationwide serosurvey in the Netherlands"

*To accompany the manuscript:*

**Table S1.** Crosstabulation of age-aggregated PICO-1 data by serostatus and symptomatic/not, for ECDC case definition (main analysis) and for 'any symptom' definition (sensitivity analysis 2) of symptomatic. Percentages are with respect to row totals.

| | ECDC case definition | | Any symptom | |
|---|---|---|---|---|
| | Symptomatic (%) | Not symptomatic (%) | Symptomatic (%) | Not symptomatic (%) |
| Seropositive | 44 (59%) | 30 (41%) | 51 (69%) | 23 (31%) |
| Seronegative | 912 (30%) | 2161 (70%) | 1527 (50%) | 1546 (50%) |
| Total | 956 (30%) | 2191 (70%) | 1578 (50%) | 1569 (50%) |

**Table S2.** Model fit (according to AIC) of model variants (all models include intercept). Selected model (with lowest AIC) in **boldface**.

| Included terms | AIC | AIC difference* |
|---|---|---|
| 1. Intercept only | 4192.0 | 46.1 |
| 2. Serostatus | 4177.5 | 21.6 |
| 3a. Serostatus + age (linear term) | 4147.1 | 1.2 |
| 3b. Serostatus + age (P-spline) | 4146.1 | 0.2 |
| **4. Serostatus × age (separate P-splines fitted for positive & negative)** | **4145.9** | – |
| 5. Serostatus × age (separate P-splines) + sex | 4147.7 | 1.8 |
| 6. Serostatus × age (separate P-splines) + educ | 4147.7 | 1.8 |
| 7. Serostatus × age (separate P-splines) + household size category | 4147.2 | 1.3 |
| 8. Serostatus × age (separate P-splines) + sex + educ + healthcare occupation | 4146.8 | 0.9 |

* Compared with selected model (4)

**Table S3.** Model fits (according to AIC) for sensitivity analysis 2. Selected model (with lowest AIC) in **boldface**.

| **Included terms** | **AIC** | **AIC difference*** |
|---|---|---|
| 1. Intercept only | 5336.6 | 39.7 |
| 2. Serostatus | 5333.8 | 36.9 |
| 3a. Serostatus + age (linear term) | 5315.4 | 18.5 |
| 3b. Serostatus + age (P-spline) | 5300.7 | 3.8 |
| 4. Serostatus × age (separate P-splines fitted for positive & negative) | 5299.9 | 3.0 |
| 5. Serostatus × age (separate P-splines) + sex | 5298.2 | 1.3 |
| **6. Serostatus × age (separate P-splines) + sex + educ** | **5296.9** | – |
| 7. Serostatus × age (separate P-splines) + sex + educ + household size category | 5298.9 | 2.0 |
| 8. Serostatus × age (separate P-splines) + sex + educ + healthcare occupation | 5298.5 | 1.6 |

* Compared with selected model (6)

**Fig. S1.** Results of sensitivity analysis 1: estimated asymptomatic proportion per 10-year age-group comparing three different assumptions regarding the size of the antibody response window (corresponding to the median number of days assumed needed to build an IgG response).

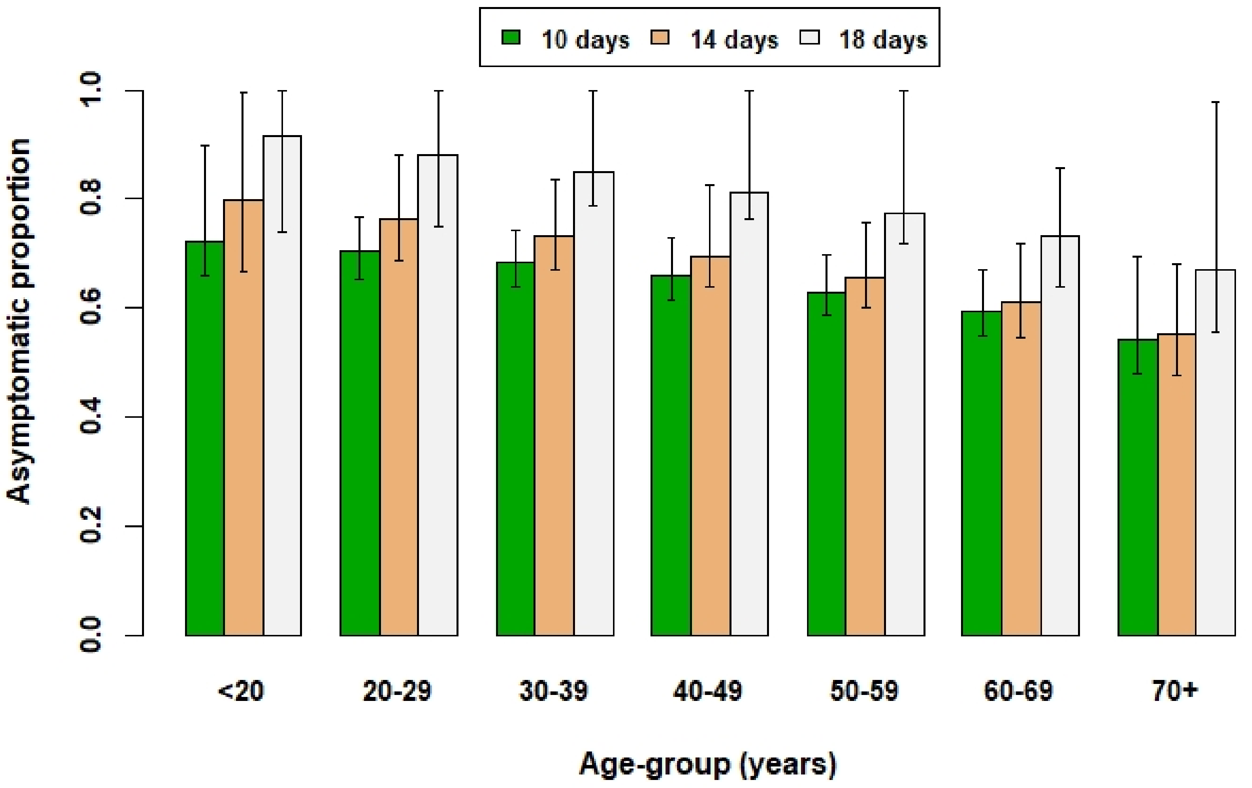

**Fig. S2.** Results of sensitivity analysis 2 ('any symptom' definition): the estimated asymptomatic proportion per 10-year age-group, as derived using the attributable risk approach.

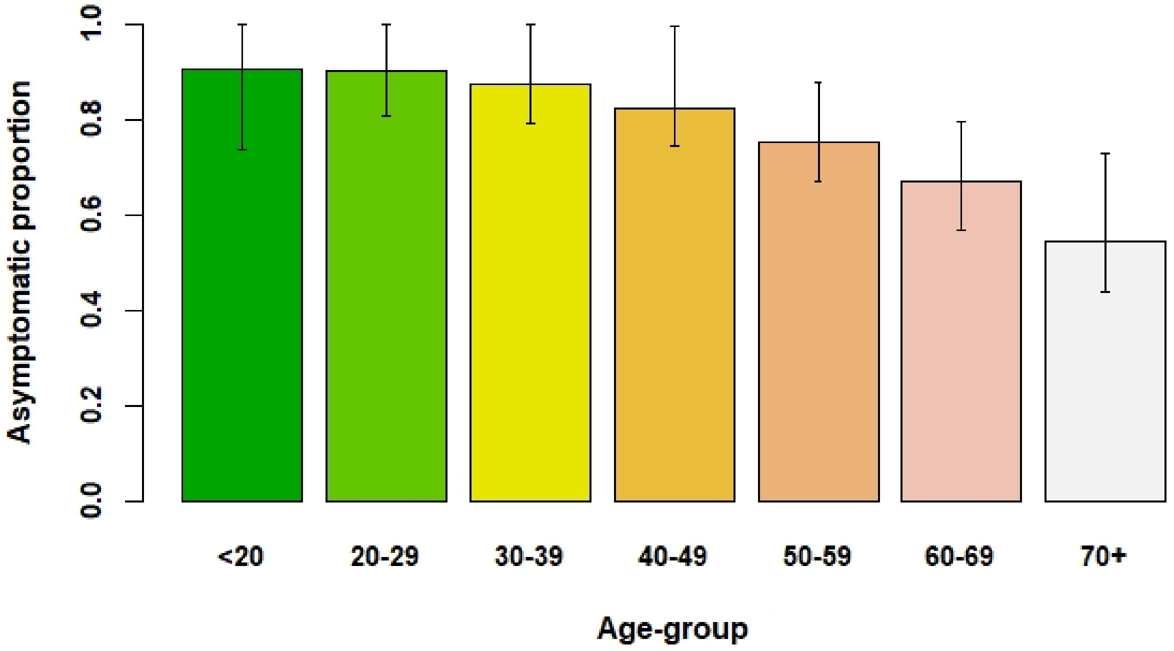
